# COVID-19 Due to Wild-Type SARS-CoV-2 More Prevalent in Adolescents and Youth than in Older Adults Based on 19 US States in Fall 2020 Prior to Vaccine Availability

**DOI:** 10.1101/2021.07.06.21260112

**Authors:** Barbara Rumain, Moshe Schneiderman, Allan Geliebter

## Abstract

**PURPOSE:** In a prior study, we examined data from six US states during Summer 2020, and found that prevalence of COVID-19 for adolescents and youth was significantly greater than for older adults (p<.00001) as was a prevalence-related measure: Number of cases observed ÷ Number of cases expected (p<.005). We now extended our study to more states in Fall 2020 to confirm the prevalence relationships we found previously. Vaccines were still not available as of Fall 2020. Presumably, the SARS-CoV-2 strain circulating at the time was the wild-type lineage since no variants were reported in the US until the end of December 2020.

**METHODS:** We examined data from 19 U.S. states experiencing surges in cases to determine prevalence of COVID-19, and a prevalence-related measure: [Number of cases observed in a given age group] ÷ [Number of cases expected in the age group based on population demographics].

**RESULTS:** In 16 of the 19 states, we found that: (1) prevalence of COVID-19 for adolescents and youth was significantly greater than for older adults (*p*-values ranged from p<0.00001 to *p* = 0.0175; (2) the ratio of cases observed to cases expected was significantly greater in adolescents and youth than in older adults (*p*-values ranging from *p*< 0.00001 to *p* = 0.004).

**CONCLUSIONS:** Our results are consistent with our previous study in Summer 2020. The finding of lower prevalence in older adults cannot be attributed to access to vaccination since our data are from Fall 2020 when vaccinations were not yet available. Our findings with the SARS-CoV-2 wild-type strain are consistent with the findings currently being reported in the UK for the delta variant. In both studies, prevalence in adolescents and youth exceeded that in older adults. The UK findings are more pronounced perhaps because that study transpired following months of vaccinations of older adults whereas ours occurred before vaccinations were available.

## INTRODUCTION

The susceptibility of adolescents (10-19) and youth (15-24) to COVID-19 has been a matter of controversy. In the very early studies conducted in China, Dong et al. [1], Lu et al. [2], and Bi *et al*. [3] reported that adolescents were susceptible, with Bi *et al*. [3] reporting that the rate of infection across all age groups was similar. However, Zhang et al. [4], in a study in Hunan province, China, concluded that older adults were the most susceptible, those in the first half of adolescence least susceptible, and youth (15–24) intermediate in susceptibility. Geliebter, Rumain & Schneiderman [5] attempted to replicate Zhang *et al*.’s statistical analyses but obtained results in line with those of Bi *et al*., indicating a similar infection rate across the age groups. Nevertheless, other data from Europe also indicated that adolescents were less susceptible than adults (Kuchar *et al*. in Warsaw [6]; de Lusignan *et al*. [7] in England). Also, Viner et al. [8], after a meta-analysis of 32 studies, concluded that “children and adolescents younger than 20 years had 44% lower odds of secondary infection with SARS-CoV-2 as compared with adults 20 years and older.” Subsequently, a mathematical model by Eggo and her colleagues [9], based on data from China, Italy, Japan, Singapore, Canada and South Korea, estimated that the susceptibility of adolescents 10-19-year-olds (mean = .38) was less than half that of older adults, ages 60+ (mean = .81), but that youth in their early 20s had a susceptibility almost equal to that of older adults. However, no U.S. data were included in their model.

We [10] therefore decided to examine data from the U.S. We calculated the prevalence in adolescents and youth compared to that of older adults in the U.S. in the summer of 2020. We analyzed the health department data from six US states that were experiencing surges in cases. In all six states, we found that: (1) prevalence of COVID-19 for adolescents and youth was significantly greater than for older adults, p < .00001, as was (2) the ratio of observed to expected cases, p < .005. Vaccines were not yet available during the time of our study, and therefore, our findings were not an artifact of older adults being vaccinated first.

One limitation in our prior study was that it was based on data from only six U.S. states. The purpose of the present study was to investigate the prevalence of COVID-19 in adolescents and youth compared to that in older adults, in 19 states that met certain criteria. We also wanted to confirm the relationships we found during Summer 2020 in the Fall of 2020, when states were experiencing new surges in cases. Vaccines were still not available as of Fall 2020.

## METHODS

There were two criteria for inclusion of states in our study sample:

1. The state experienced a surge during Fall 2020, defined as follows: After at least a 1-month plateau in the 7-day average of daily number of new cases, there was a dramatic increase of at least 75% from the plateau 2–3 months prior, which lasted at least one month, as reported for the states in the New York Times “COVID Map and Case Count” [11]. As an example, for Colorado, the case data are from October 22^nd^ when there was a surge. On that day, the 7-day average number of daily new cases was 1,171. Sixty days prior on August 22^nd^, the 7-day daily average was 296, representing a 300% increase over August 22^nd^, and the surge was at least one month in duration. For the months of June, July, and through August 21, the 7-day daily average of new cases had plateaued at 200– 500 cases per day. Another example is Wisconsin, where the case data are from October 23^rd^. On that day the7-day daily average was 3,547. The surge lasted at least a month from October 23^rd^, with November 23^rd^ reporting a 7-day average of daily new cases at 6,436. Sixty days prior, on August 23^rd^, the 7-day daily average of new cases was 708. Thus, there was > 400% increase in the 7-day daily average. Prior to that, for the months of June, July and August, the 7-day daily average of new cases had reached a plateau, fluctuating from 300–962 cases per day. Even from the maximum of 962, the 3,547 cases on October 23^rd^ represented a 268% increase.
2. The pediatric data were tabulated within distinct age brackets, not amalgamated. California lumped all child data 0-17 years of age together, and could not be included as children under age 10 are excluded. Massachusetts lumped children 0-19 together, as did Arizona. We therefore considered the following 19 states: Alabama, Alaska, Colorado, South Carolina, New Mexico, Michigan, Montana, Wisconsin, Rhode Island, South Dakota, Missouri, Oklahoma, North Dakota, Oregon, Nevada, Pennsylvania, Minnesota, Florida, and Tennessee. We accessed online tables containing COVID-19 case data when there was a surge from state Health Department websites, and tables for state population data by age group. Case data were downloaded between October 22^nd^ and December 5th, at the time each of the states was experiencing a spike in cases. The websites are detailed in S1 Appendix, which includes the relevant tables/figures.

Depending on how the data were tabulated, the case data for the 19 states were either for adolescents (ages 10-19), for youth (ages 15-24), or for adolescents and youth combined.

### Adolescents

In the following 13 states, data were tabulated by decade, and we examined the 10-19-year age bracket: Alaska, Colorado, New Mexico, Michigan, Wisconsin, Montana, South Dakota, North Dakota, Oregon, Nevada, and Pennsylvania. Tennessee had a similar age bracket of 11–20 years of age. Thus these 13 states provided data on adolescents.

### Youth

The four states of Florida, Oklahoma, Rhode Island and Alabama, provided the data on youth. The age brackets for Florida and for Oklahoma were 5–14, and 15–24 (“youth” as defined by the WHO). Therefore, we used only the 15-24-year-olds for both these states. We also used the 15-24 age bracket for Rhode Island. For Alabama, cases were reported for 0-4, 5-17 years and for 18–24 years--age bracket demarcations unlike the other states, and we used the 18-24 years age bracket, since this is a subset of the “youth” age bracket.

### Adolescents and Youth Combined

Minnesota and Missouri provided data on adolescents and youth combined. For both Minnesota and Missouri, we considered the age bracket 10-24-year-olds.

The case data from the Health Department websites was used to compute “Percentage of Cases Observed,” calculated as the number of cases in a particular age group divided by the total number of cases for all ages in the state and converted into a percentage. The “Percentage of Cases Expected” was determined based on population demographics: For each age group, it was the percentage of the population the given age group comprises, multiplied by the total number of cases. The population demographic data were obtained from the state websites, listed in S1 Appendix.

We then calculated two measures: 1) Prevalence, and 2) “Percentage of Cases Observed” in a given age group ÷ “Percentage of Cases Expected” based on population demographics as noted above. For an illustration of how these measures were calculated, see Rumain et al. (2021). Statistics: We performed chi-square calculations to determine whether differences between the adolescent/youth groups and the older adults were significant for the two outcome measures. Significance level was based on 2-tailed α = .05.

## RESULTS

In 16 of the 19 states, we found that: (1) prevalence of COVID-19 for adolescents and youth was significantly greater than for older adults, with *p*-values varying from p<0.00001 to *p* = 0.0175 ; and (2) the ratio of observed number of cases to expected number of cases was significantly greater in adolescents and youth than in older adults, with *p*-values varying from *p*< 0.00001 to *p* = 0.004, (**Table 1**), with more cases than one would expect based on the demographics of the populations. The three states that did not follow the general pattern were: South Dakota, Michigan and Pennsylvania.

**Table 1.**
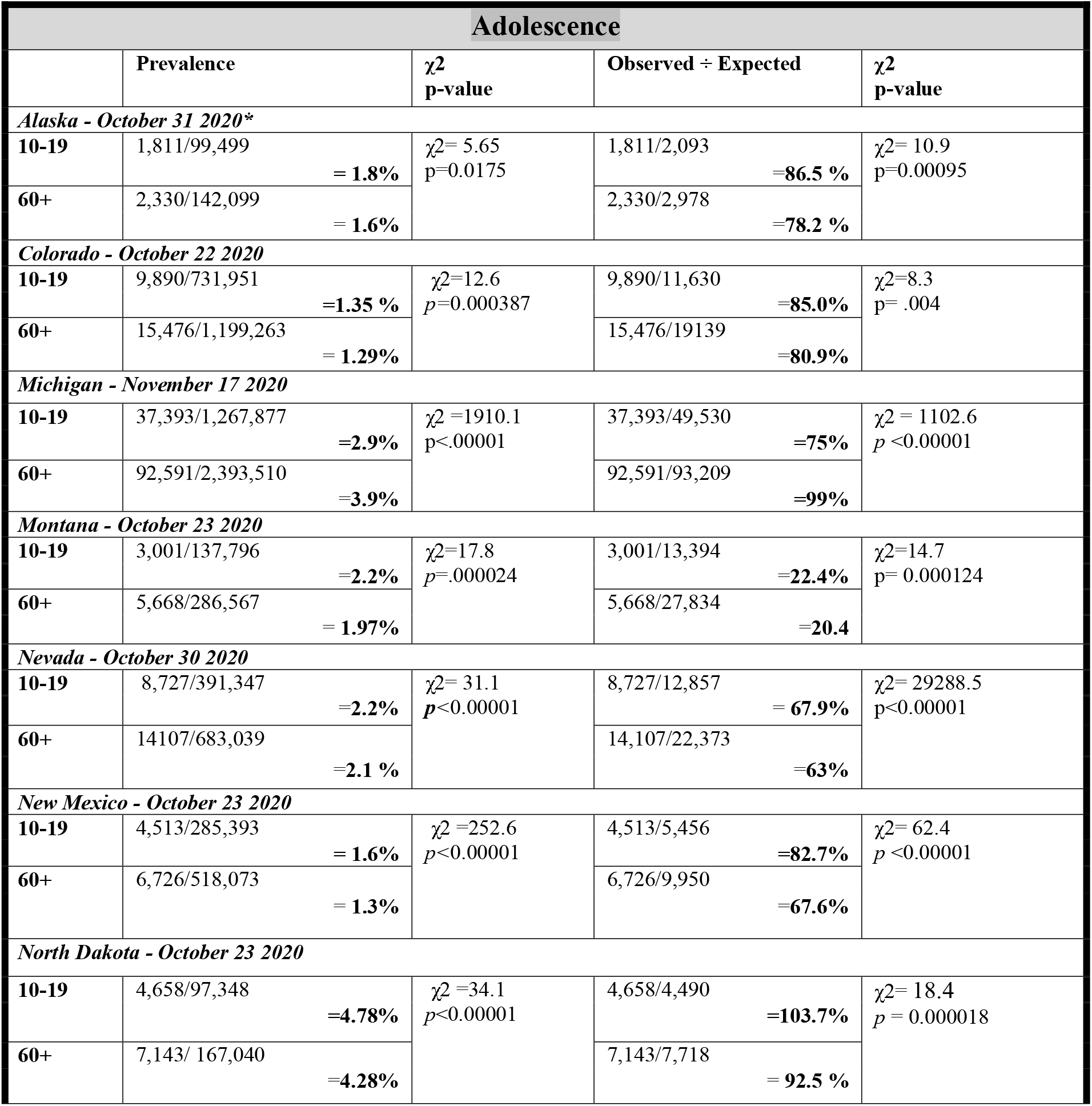
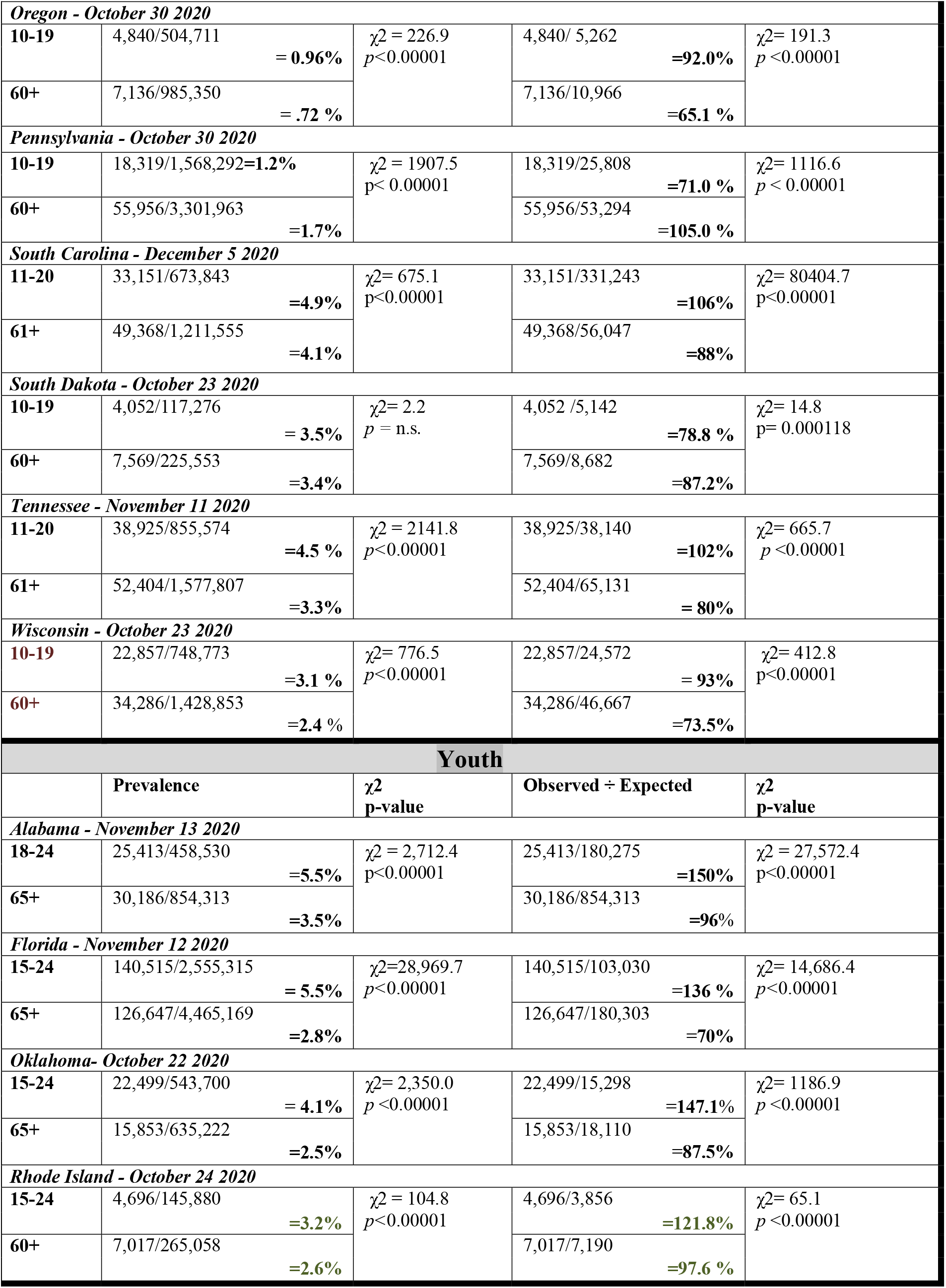
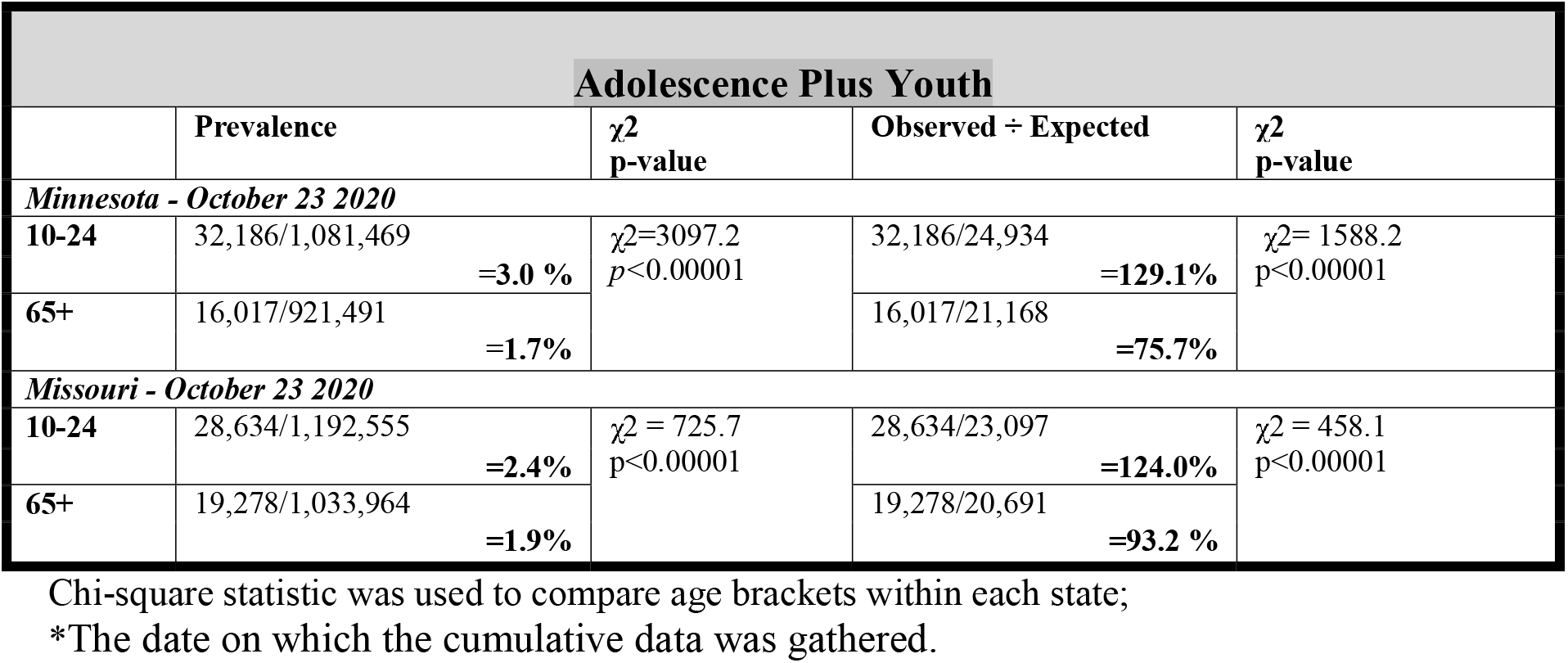
Prevalence-related Outcome Measures by Developmental Period and Age Bracket in US States Experiencing Spikes in COVID-19 Cases.

We now consider each developmental period separately: Adolescence, Youth, and Adolescence plus Youth Combined.

### Adolescence (10-19-year-olds): Data from 13 States: Alaska, Colorado, Michigan, Montana, Nevada, New Mexico, North Dakota, Oregon, Pennsylvania, South Carolina, South Dakota, Tennessee and Wisconsin

#### Prevalence

The prevalence in adolescents was significantly greater than that in older adults in 10 of the 13 states (Alaska, p=.02; Colorado, p=0.004; Montana, p=0.0002; Nevada, New Mexico, North Dakota, Oregon, South Carolina, Tennessee, and Wisconsin, p< 0.00001). For South Dakota, there was no significant difference between the adolescents and older adults. For Michigan and Pennsylvania, the pattern was reversed with the prevalence in older adults being significantly higher than that in adolescents (p< 0.00001).

#### Proportion of cases observed to cases expected

The ratio of the number of cases observed to the number of cases expected based on population demographics, was significantly greater for adolescents than for older adults for 10 of the 13 states: Alaska, p = 0.00095; Colorado, p= 0.004; Montana, p = 0.000124; North Dakota, p= 0.00002; Nevada, New Mexico, Oregon, South Carolina, Tennessee and Wisconsin, all p < 0.00001. For Michigan, Pennsylvania, and South Dakota, the pattern was reversed with the proportion of cases observed to cases expected being significantly greater in older adults than in adolescents (for the former two states, p<0.00001, and for the latter one, p = 0.000118).

### Youth (15-24-year-olds): Data from 4 States: Alabama, Florida, Oklahoma, Rhode Island

#### Prevalence

In Alabama, the prevalence in youth 18-24-years-old was 157% that of older adults (5.5% vs. 3.5%, p<0.00001). In Florida, the prevalence in youth 15-24 years of age was 196% that of older adults, p<0.00001; in Oklahoma, it was 164% that of older adults, p<0.00001, and in Rhode Island, it was 123% that of older adults, p<0.00001.

#### Proportion of cases observed to cases expected

In Alabama, the ratio of observed to expected cases was approximately in 18–24-year-olds 150% that of older adults (150% vs. 96%), p < 0.00001; in Florida, the ratio in youth was almost 200%t that of older adults (136% v. 70%), p < 0.00001). In Oklahoma, the ratio of observed to expected cases for youth was 167% that of older adults (147.1% vs. 87.5%), p < 0.00001. And, in Rhode Island, it was 125% that of older adults (121.8% vs. 97.6%), p<0.00001.

### Adolescence plus Youth Combined (ages 10–24 years-old): Data from 2 States: Minnesota and Missouri

In both Minnesota and Missouri, the prevalence of COVID-19 in 10–24-year-olds was significantly greater than it was in older adults, > 65 years (p<0.00001). Also, in both states, the proportion of observed cases of COVID-19 to expected cases based was significantly greater for the 10-24-year-olds than it was for the older adults, p<0.00001.

In 16 of the 19 states experiencing surges of cases during Fall 2020, the prevalence of COVID-19 was significantly higher in adolescents and youth than it was in older adults, p = 0.0022, sign test.

## DISCUSSION

In 16 of the 19 states experiencing surges of cases during Fall 2020, we found that (i) the prevalence of COVID-19 was significantly higher in adolescents and youth than in older adults, and (ii) similarly the ratio of observed cases of COVID-19 to expected cases based on population demographics was significantly greater (p<0.00001). Our data is presumably on the original wild type SARS-CoV-2 strain (i.e., the strain with no major mutations) since as of October and November 2020, there were no reports of variants in the US: The first variant reported in the US, the B.1.1.7, was reported at the end of December 2020 [12]. All the data are from October - November 2020 except for South Carolina which is from December 5, 2020—all before the report of any variants in the U.S. Moreover, our findings of lower prevalence in older adults cannot be attributed to access to vaccination since our data are from Fall 2020 when vaccinations were not yet available. A possible reason for our findings is that adolescents have more contacts than adults [13], and another factor, is that older adults, feeling vulnerable, may be more likely to adhere to masking and social distancing, which adolescents/youth may disregard. Both these factors are likely operative.

Our findings in the six U.S. states differ from those of Zhang et al. in China, who found that the infection rate in older adults, ages 65+, exceeded that in adolescents and youth, and from those of Wu et al. [14], who found that of 44,672 confirmed cases of COVID in mainland China, only 1% were in adolescents ages 10-19 years of age. Out findings also contradict Davies et al.’s model that estimates 10-19-year-olds’ susceptibility to be half that of older adults. The reason for the discrepancy could be these earlier studies were conducted when schools were closed, which reduced the number of contacts by adolescents and youth, and thus the number of cases. Moreover, testing was not readily available early on in the pandemic, and adolescents tend to have milder cases of COVID-19 that could have been missed without the availability of widespread testing. In line with this reasoning is as of April 2, 2020, among 149,082 cases in all age groups for which patient age was known, only 2,572 (1.7%) of these occurred in children aged <18 years, with nearly 60% of these cases occurring in adolescents 10-17 years old [15]. Hence at that point, adolescents accounted for just 1% of the total cases. But by Sept 15, 2020, a month or two before our data was collected, the number of cases in adolescents 10-19 years of age had climbed to 387,000 [16]. And, as of June 24, 2021, the American Academy of Pediatrics reported 4,032,782 total confirmed COVID-19 cases in children <18 years old [17] with at least 336 deaths.

We now consider the three states that were an exception and whether events occurring in these states might explain the difference in findings. There were two types of events occurring.

The first such event, the Sturgis Motorcycle Rally held annually in South Dakota, easily explains the reversal from the usual pattern that we find in South Dakota. The data from South Dakota were from October 23, 2020, and followed the Sturgis Motorcycle Rally where half a million bikers converged on Sturgis between August 7 and August 16, 2020. Dave et al. [18] link a deluge of COVID-19 cases to this event since social distancing and mask-wearing were rare in Sturgis [19, 20]. The event led to increased cases in South Dakota as well as in other states from where bikers travelled to South Dakota. Using anonymized cell phone data from SafeGraph Inc., the authors traced smartphone pings from non-residents and indicating that visitors came to Sturgis from all parts of the US, including states that recently experienced surges in COVID-19 cases. The attendees opened themselves up to infection at this super-spreader event, and then contributed to increased case counts in their home states when they returned home. These attendees were mostly adults who would be more likely to come into contact with other adults rather than adolescents and youth, thereby increasing the number of cases in adults, including older adults.

This explanation is bolstered by prior data on South Dakota from Summer 2020, i.e., from September 4^th^, when the prevalence of COVID-19 in 10–19-year-olds was significantly greater than it was in > 60-year-olds (p<0.00001). Similarly, the proportion of cases observed to expected, was significantly greater in 10-19-year-olds than in those 60+ (p<0.00001). Six weeks later, as more cases from the Sturgis event developed, there was no longer a significant difference in the prevalence between the adolescents and older adults.

As for Pennsylvania and Michigan, the operative event that may have led to a reversal in the expected pattern of results was the Trump rallies. Both Pennsylvania and Michigan were important for Trump to win the election and were the sites of many Trump rallies, which were known to be largely without masking and social distancing. In Pennsylvania, the data are from October 30. From August through October 30^th^, between Trump and his son Don Jr., there were 15 Trump rallies held in Pennsylvania. Trump held nine of these rallies in Pennsylvania [21]: Old Forge, Pa. (August 20, 2020); Latrobe, Pa. (September 3, 2020); Moon Township, Pa. (September 22, 2020); Somerset County Sept 11, 2020); Middletown, Pa. (September 26); Johnston, Pa. (October 13, 2020); Erie, Pa. (October 20, 2020); Allentown, Pa. (October 26, 2020); Lititz, Pa. (October 26, 2020); and Martinsburg, Pa. (October 26, 2020). Don Jr. held an additional 6 of the Pennsylvania rallies [22-26] at the following locations on the following dates: Blue Ridge Sportsman Club outside Harrisburg, Pa. (Sept. 16, 2020); Roxbury Park Bandshell, Johnstown, Pa. (September 23, 2020); Nittany Valley/Centre County, Pa. (October 19, 2020); Blair County, Pa. (October 19, 2020); York Springs, Pa. (October 30, 2020); Ambridge, Pa. (October 30, 2020).

In Michigan, the data are from November 17, 2020, and in the months leading up to the election, Trump and his son Don Jr. held 11 rallies in the state [21]. They were: Freeland, MI (September 10, 2020), Muskegon, MI (October 17, 2020), Macomb County (Don Jr., on October 26), Lansing, MI (October 27, 2020), Waterford Township, MI (October 30, 2020), Davison, MI (Don Jr., October 31, 2020), Grand Bay Marine in Traverse City (Don Jr., October 31, 2020), Macomb County (November 1, 2020), Washington, MI (November 1, 2020), Traverse City, MI (November 2, 2020), Grand Rapids, MI (November 2, 2020). These rallies attracted adults, including older adults, rather than adolescents or youth, and with the lack of social distancing and mask wearing, likely added to the caseloads of older adults. Whereas there were also Trump rallies in some of the other sates we examined, there were far fewer of them: There were 6 in Florida, 1 in Oklahoma, 3 in Minnesota, and 6 in Wisconsin [21] by the dates for the COVID— 19 tabulated state data, which are 11/12/2020, 10/22/2020, 10/23/2020, and 10/23/2020, respectively. The confluence of the two factors noted—the Sturgis motorcycle super-spreader event and the Trump rallies—likely inflated the number of cases in the older adults.

Interestingly, our finding of higher prevalence in adolescents and youth for the wild-type strain parallels the recent findings of the REACT-1 (REal-time Assessment of Community Transmission-1) UK study at Imperial College London for the delta variant. The findings indicate a five-fold higher positivity rate among 5–12-year-olds and youth 18-24 than among older adults, ages 65+ [27]. The findings are being interpreted to mean that youth are driving the UK surge with regards to the delta variant [28]. However, our findings indicate that the high prevalence in adolescents and youth is not a novel phenomenon and was present with the wild-type strain in our previous study and in the current one. One reason the UK study found a five-fold difference in positivity which is greater than ours may be that their study was conducted after vaccinations had been ongoing in the UK for many months for older adults, which would drive down the positivity in that cohort. Our study in the US was conducted prior to the availability of vaccinations, and may be the reason the difference in prevalence between the adolescents/youth vs. older adults was less pronounced.

Our finding of the high prevalence of COVID-19 in adolescents and youth in the U.S., taken together with the high virulence and high transmissibility of the delta variant currently circulating in the US, should be considered in any decision about masking in these groups, especially in those under 12, for whom vaccinations are not yet available.

## CONCLUSIONS

The findings of high prevalence of COVID-19 and the other prevalence-related measure in adolescents and youth in the U.S. are consistent with our earlier findings in the Summer of 2020. Our findings for the wild-type strain of SARS-CoV-2, are also consistent with the findings of the ongoing REACT-1 study being conducted in the UK with the delta variant. The disparity in prevalence occurs even with the wild-type SARS-CoV-2 lineage, although the disparity between the age groups is not as pronounced. The lower disparity may be the result of our study occurring before vaccinations were available, whereas the UK study took place after months of vaccinating older adults.

## Data Availability

Data Availability Statement:  The Supporting Information file, S1 Appendix, contains a listing of websites and URLs where the publicly available research data that we relied on, can be found. It also contains the actual tables and figures we used. 

## Notes

### Competing Interest Statement

The authors have declared no competing interest.

### Funding Statement

There was no source of funding for the study.

### Author Declarations

The study is exempt because it uses previously collected aggregated health data collected by state health departments.

