## Supplementary material for "COVID-19 Due to Wild-Type SARS-CoV-2 More Prevalent in Adolescents and Youth than in Older Adults Based on 19 US States in Fall 2020 Prior to Vaccine Availability": S1 Appendix Supplement

Appendix S1, Supplemental material for the manuscript:

"COVID-19 Due to Wild-Type SARS-CoV-2 More Prevalent in Adolescents and Youth than in Older Adults Based on 19 US States in Fall 2020 Prior to Vaccine Availability"

by B. Rumain, M. Schneiderman, A. Geliebter

**Methodological Details**

This supplementary material is provided by the authors to give readers additional source information about the data used in this study.

#### S1.1. Case Data by State Websites:

(i)Alaska: https://experience.arcgis.com/experience/2d19dc2b5c7e4b399ff6495a8950493d/

[Retrieved Nov. 1, 2020]

(ii) Colorado: https://covid19.colorado.gov/data [Retrieved Oct. 23, 2020]

(iii) Michigan: https://www.mistartmap.info/compare/age-group [Retrieved Nov. 18, 2020]

(iv) Montana: https://dphhs.mt.gov/publichealth/cdepi/diseases/coronavirusmt/demographics

[Retrieved Oct. 24, 2020]

(v) Nevada: https://nvhealthresponse.nv.gov [Retrieved Oct. 31, 2020]

(vi) New Mexico: https://cvprovider.nmhealth.org/public-dashboard.html# [Retrieved Oct. 24, 2020]

(vii) North Dakota: https://www.health.nd.gov/diseases-conditions/coronavirus/north-dakota-coronavirus-cases [Retrieved October 24, 2020]

(viii) Oregon:
https://public.tableau.com/app/profile/oregon.health.authority.covid.19/viz/OregonCOVID-19CaseDemographicsandDiseaseSeverityStatewide/DemographicData [Retrieved Oct. 31, 2020]

(ix) Pennsylvania: https://experience.arcgis.com/experience/ed2def13f9b045eda9f7d22dbc9b500e
[Retrieved Oct. 31, 2020]

(x) South Carolina: https://scdhec.gov/covid19/south-carolina-county-level-data-covid-19
[Retrieved May 15, 2021]

(xi) South Dakota: https://doh.sd.gov/news/coronavirus.aspx [Retrieved Oct. 24, 2020]

(xii) Tennessee: https://www.tn.gov/content/tn/health/cedep/ncov/data.html [Retrieved Nov. 12, 2020]

(xiii) Wisconsin: https://www.dhs.wisconsin.gov/covid-19/cases.htm#by%20age [Retrieved Oct. 24, 2020]

(xiv) Alabama: https://bamatracker.com/chart/caseages [Retrieved Nov. 14, 2020]

(xv) Florida: https://floridahealthcovid19.gov/ [Retrieved November 13, 2020]

(xvi)Oklahoma:https://coronavirus.health.ok.gov/sites/g/files/gmc786/f/2020.10.23_weekly_epi_report.pdf [Retrieved Oct. 23, 2020]

(xvii) Rhode Island: https://ri-department-of-health-covid-19-case-data-rihealth.hub.arcgis.com/#age [Retrieved October 25, 2020]

(xviii) Minnesota: https://www.health.state.mn.us/diseases/coronavirus/situation.html#ageg1 [Retrieved Oct. 24, 2020]

(xix) Missouri: https://health.mo.gov/living/healthcondiseases/communicable/novel-coronavirus/data/public-health/demographics.php [Retrieved Oct. 24, 2020]

**S1.2. Demographic Data by Age and by State websites:**

(i) Alaska: https://live.laborstats.alaska.gov/pop/

https://demography.dola.colorado.gov/population/data/sya-regions/

(ii) Colorado: Colorado State Demography Office, Colorado Department of Local Affairs,

https://demography.dola.colorado.gov/population/data/sya-regions/

(iii) Michigan: https://censusreporter.org/profiles/04000US26-michigan/

(iv) Montana: https://censusreporter.org/profiles/04000US30-montana/

(v) Nevada: https://censusreporter.org/profiles/04000US32-nevada/

(vi) New Mexico: https://censusreporter.org/data/table/?table=B01001&geo_ids=04000US35&primary_geo_id=04000US35

(vii) North Dakota: https://censusreporter.org/profiles/04000US38-north-dakota/

(viii) Oregon: https://censusreporter.org/profiles/04000US41-oregon/

(ix) Pennsylvania: https://censusreporter.org/profiles/04000US42-pennsylvania/

(x) South Carolina https://censusreporter.org/profiles/04000US45-south-carolina/

(xi) South Dakota: https://www.sdstate.edu/sociology-rural-studies/census-data-center/population-change

(xii) Tennessee: https://www.tn.gov/content/dam/tn/health/documents/population/TN-Population-by-AgeGrp-Sex-Race-Ethnicity-2019.pdf

(xiii) Wisconsin: https://censusreporter.org/profiles/04000US55-wisconsin/

(xiv) Alabama: https://censusreporter.org/profiles/04000US01-alabama/

(xv) Florida: http://edr.state.fl.us/Content/populationdemographics/data/Pop_Census_Day.pdf

(xvi) Oklahoma: https://censusreporter.org/profiles/04000US40-oklahoma/

(xvii) Rhode Island: https://censusreporter.org/profiles/04000US44-rhode-island/

(xviii) Minnesota: https://censusreporter.org/profiles/04000US27-minnesota/

(xix) Missouri: https://healthapps.dhss.mo.gov/MoPhims/QueryBuilder?qbc=PNM&q=1&m=1

**S1.3. New York Times Websites Indicating Surges by State:**

https://www.nytimes.com/interactive/2021/us/alaska-covid-cases.html

https://www.nytimes.com/interactive/2021/us/colorado-covid-cases.html

https://www.nytimes.com/interactive/2021/us/michigan-covid-cases.html

https://www.nytimes.com/interactive/2021/us/montana-covid-cases.html

https://www.nytimes.com/interactive/2021/us/nevada-covid-cases.html

https://www.nytimes.com/interactive/2021/us/new-mexico-covid-cases.html

https://www.nytimes.com/interactive/2021/us/north-dakota-covid-cases.html

https://www.nytimes.com/interactive/2021/us/oregon-covid-cases.html

https://www.nytimes.com/interactive/2021/us/pennsylvania-covid-cases.html

https://www.nytimes.com/interactive/2021/us/south-carolina-covid-cases.html

https://www.nytimes.com/interactive/2020/us/south-dakota-coronavirus-cases.html

https://www.nytimes.com/interactive/2020/us/tennessee-coronavirus-cases.html

https://www.nytimes.com/interactive/2021/us/wisconsin-covid-cases.html

https://www.nytimes.com/interactive/2021/us/alabama-covid-cases.html

https://www.nytimes.com/interactive/2020/us/florida-coronavirus-cases.html

https://www.nytimes.com/interactive/2021/us/oklahoma-covid-cases.html

https://www.nytimes.com/interactive/2021/us/rhode-island-covid-cases.html

https://www.nytimes.com/interactive/2021/us/minnesota-covid-cases.html

https://www.nytimes.com/interactive/2020/us/missouri-coronavirus-cases.html

Since the data on the websites above are updated on a daily basis, we are including below in S1.4 the tables of the data the way they appeared on the websites when we accessed them. These provide the numbers we used in our calculations.

**S1.4. List of Supplemental Tables.**

1) Table A. Alaska COVID-19 Cases by Age (as of October 31, 2020)
2) Table B. Colorado COVID-19 Cases by Age (as of Oct. 22, 2020)
3) Table C. Michigan COVID-19 Cases by Age (as of Nov. 17, 2020)
4) Table D. Montana COVID-19 Cases by Age (as of Oct. 23, 2020)
5) Table E. Nevada COVID-19 Cases by Age (as of October 30, 2020)
6) Table F. New Mexico COVID-19 Cases by Age (as of Oct. 23, 2020)
7) Table G. North Dakota COVID-19 Cases by Age (as of October 23, 2020)
8) Table H. Oregon COVID-19 Cases by Age (as of October 30. 2020)
9) Table I. Pennsylvania COVID-19 Cases by Age (as of October 30, 2020)
10) Table J. South Carolina COVID-19 Cases by Age (as of Dec. 5, 2020)
11) Table K. South Dakota COVID-19 Cases by Age (as of Oct. 23, 2020)
12) Table L. Tennessee COVID-19 Cases by Age (as of November 11, 2020)
13) Table M. Wisconsin COVID-19 Cases by Age (as of Oct. 23, 2020)
14) Table N. Alabama COVID-19 Cases by Age (as of November 13, 2020)
15) Table O. Florida COVID-19 Cases by Age (as of Nov. 12, 2020)

16) Table P. Oklahoma COVID-19 Cases by Age (as of Oct. 22, 2020)

17) Table Q.   Rhode Island COVID-19 Cases by Age (as of October 24, 2020)

18) Table R.  Minnesota COVID-19 Cases by Age (as of Oct. 23, 2020)

19) Table S.  Missouri COVID-19 Cases by Age (as of Oct. 23, 2020)

Appendix S1, Supplement to:  COVID-19 Due to Wild-Type SARS-CoV-2

**Table A.  Alaska COVID-19 Cases by Age (as of October 31, 2020)**

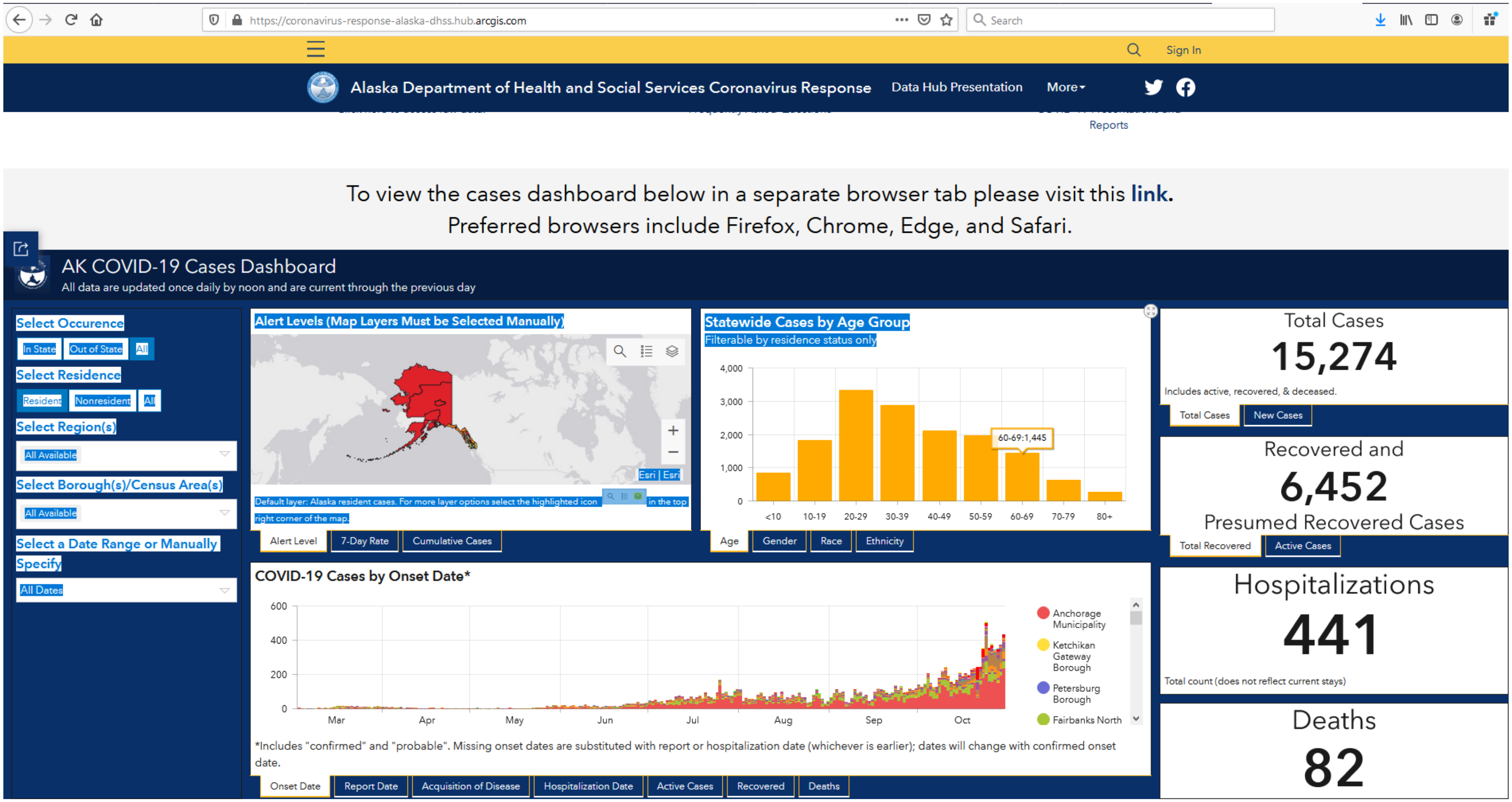


Source:  Alaska Department of Health and Social Services Coronavirus Response Hub, AK COVID—19 Cases Dashboard https://experience.arcgis.com/experience/2d19dc2b5c7e4b399ff6495a8950493d/

**Table B. Colorado COVID-19 Cases by Age as of 10/22/2020**

| Age | Proportion of Cases | Total Case Count |
|---|---|---|
| 0-9 | 0.038 | 3480 |
| 10-19 | 0.108 | 9890 |
| 20-29 | 0.223 | 20421 |
| 30-39 | 0.175 | 16025 |
| 40-49 | 0.154 | 14102 |
| 50-59 | 0.133 | 12179 |
| 60-69 | 0.085 | 7784 |
| 70-79 | 0.046 | 4212 |
| 80+ | 0.038 | 3480 |

Total cumulative cases: 91,572

Source: Colorado Department of Public Health and Environment, "CDPHE COVID19 State-Level Expanded Case Data" (as of 10/22/2020)
https://covid19.colorado.gov/data

#### Table C. Michigan COVID-19 Cases by Age (as of Nov. 17, 2020)

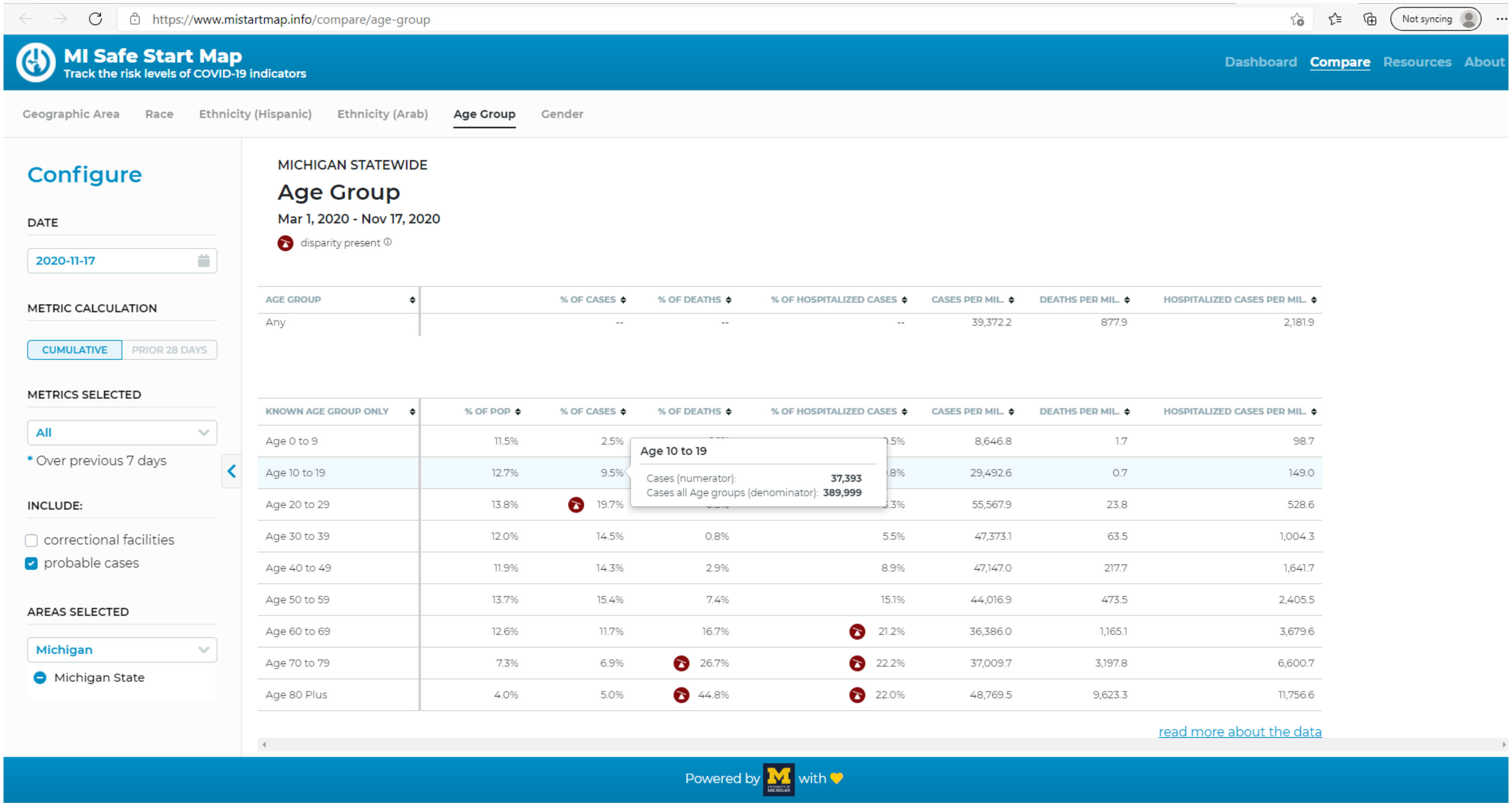


| AGE GROUP | % OF CASES | % OF DEATHS | % OF HOSPITALIZED CASES | CASES PER MIL. | DEATHS PER MIL. | HOSPITALIZED CASES PER MIL. |
|---|---|---|---|---|---|---|
| Any | -- | -- | -- | 39,372.2 | 877.9 | 2,181.9 |

| KNOWN AGE GROUP ONLY | % OF POP | % OF CASES | % OF DEATHS | % OF HOSPITALIZED CASES | CASES PER MIL. | DEATHS PER MIL. | HOSPITALIZED CASES PER MIL. |
|---|---|---|---|---|---|---|---|
| Age 0 to 9 | 11.5% | 2.5% | [illegible] | [illegible] | 8,646.8 | 1.7 | 98.7 |
| Age 10 to 19 | 12.7% | 9.5% | [illegible] | [illegible] | 29,492.6 | 0.7 | 149.0 |
| Age 20 to 29 | 13.8% | 19.7% | [illegible] | [illegible] | 55,567.9 | 23.8 | 528.6 |
| Age 30 to 39 | 12.0% | 14.5% | 0.8% | 5.5% | 47,373.1 | 63.5 | 1,004.3 |
| Age 40 to 49 | 11.9% | 14.3% | 2.9% | 8.9% | 47,147.0 | 217.7 | 1,641.7 |
| Age 50 to 59 | 13.7% | 15.4% | 7.4% | 15.1% | 44,016.9 | 473.5 | 2,405.5 |
| Age 60 to 69 | 12.6% | 11.7% | 16.7% | 21.2% | 36,386.0 | 1,165.1 | 3,679.6 |
| Age 70 to 79 | 7.3% | 6.9% | 26.7% | 22.2% | 37,009.7 | 3,197.8 | 6,600.7 |
| Age 80 Plus | 4.0% | 5.0% | 44.8% | 22.0% | 48,769.5 | 9,623.3 | 11,756.6 |

Source: MI Safe Start Map, University of Michigan

https://www.mistartmap.info/compare/age-group

**Table D. Montana COVID-19 Cases by Age (as of 10/23/2020)**

| Age Group | Number of Cases (percent of total) |
|---|---|
| 0-9 years | 1,148 (4%) |
| 10-19 years | 3,001 (11%) |
| 20-29 years | 5,452 (21%) |
| 30-39 years | 4,263 (16%) |
| 40-49 years | 3,514 (13%) |
| 50-59 years | 3,457 (13%) |
| 60-69 years | 2,883 (11%) |
| 70-79 years | 1,714 (6%) |
| 80-89 years | 796 (3%) |
| 90-99 years | 257 (1%) |
| 100+ | 18 (<1%) |
| Total Cases: | 26,503 |

Source: Montana Department of Public Health and Human Services
https://dphhs.mt.gov/publichealth/cdepi/diseases/coronavirusmt/demographics

**Table E. Nevada COVID-19 Cases by Age (as of October 30, 2020)**

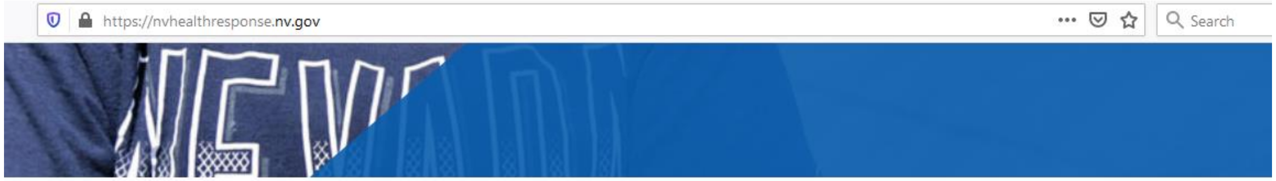


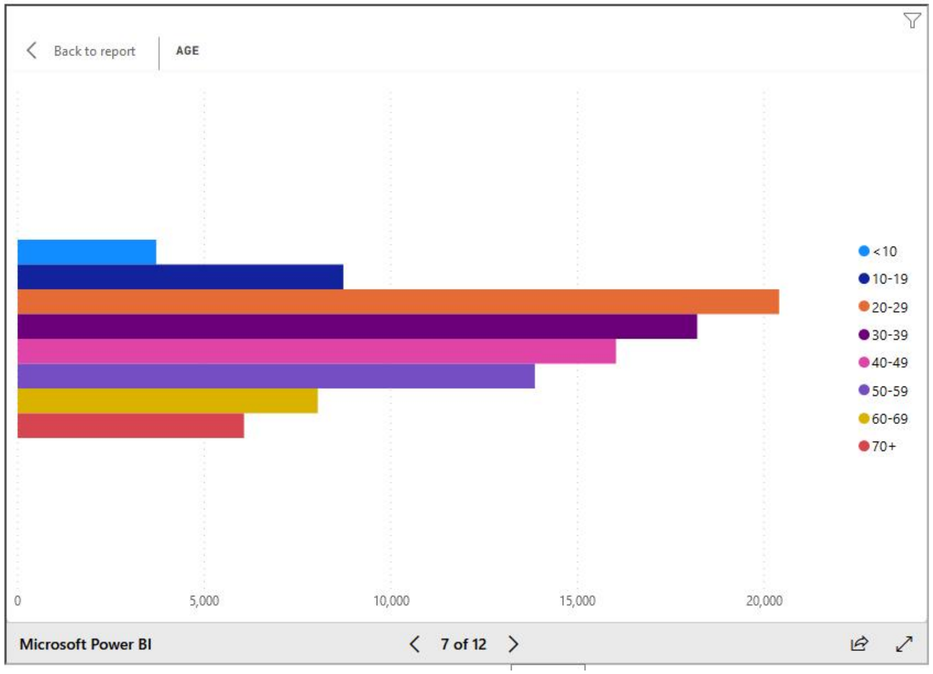


Total cases: 101,235

Source: Nevada Department of Health
[https://nvhealthresponse.nv.gov](https://nvhealthresponse.nv.gov)

The comparison of interest is between the 10-19-year-olds (8727 cases) and the older adults (60-69-year-olds: 8042 cases; 70+ year olds: 6065 cases).

**Table F. New Mexico COVID-19 Cases by Age (as of 10/23/2020)**

**Statewide Age Breakdown**

| Age | Number of Cases |
|---|---|
| 0-9 | 2120 |
| 10-19 | 4513 |
| 20-29 | 7987 |
| 30-39 | 7290 |
| 40-49 | 6145 |
| 50-59 | 5339 |
| 60-69 | 3594 |
| 70-79 | 1810 |
| 80-89 | 975 |
| 90+ | 347 |

Total: 4012

Source: New Mexico Department of Health
https://cvprovider.nmhealth.org/public-dashboard.html#

**Table G. North Dakota COVID-19 Cases by Age (as of October 23, 2020)**

| Age | Total Cases |
|---|---|
| 0-9 | 1549 |
| 10-19 | 4658 |
| 20-29 | 8814 |
| 30-39 | 5932 |
| 40-49 | 4486 |
| 50-59 | 4292 |
| 60-69 | 3490 |
| 70-79 | 1859 |
| 80+ | 1794 |
| Total | 35,080 |

Source: North Dakota Department of Health
https://www.health.nd.gov/diseases-conditions/coronavirus/north-dakota-coronavirus-cases

#### Table H. Oregon COVID-19 Cases by Age (as of October 30. 2020)

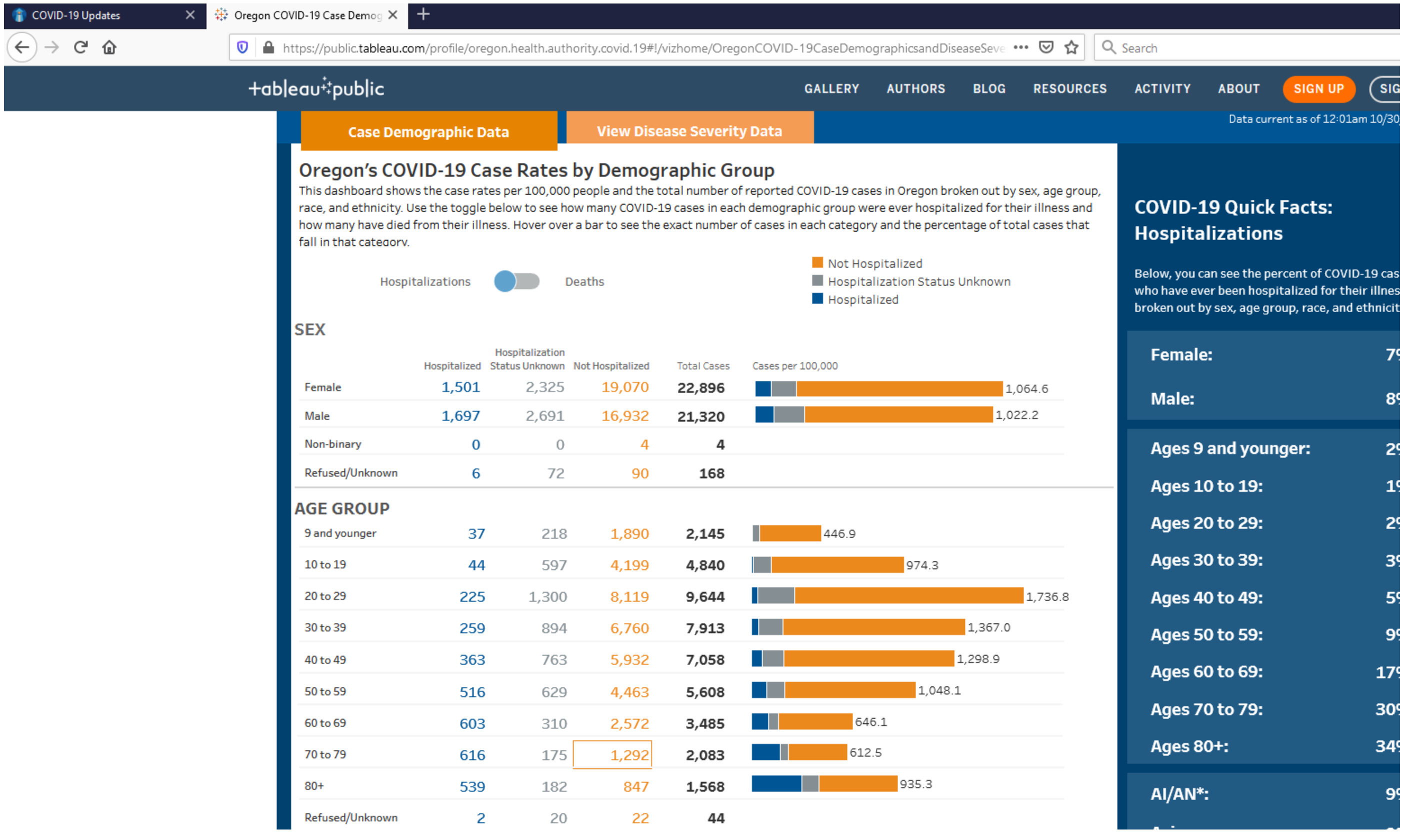

Case Demographic Data | View Disease Severity Data

Data current as of 12:01am 10/30

##### Oregon's COVID-19 Case Rates by Demographic Group

This dashboard shows the case rates per 100,000 people and the total number of reported COVID-19 cases in Oregon broken out by sex, age group, race, and ethnicity. Use the toggle below to see how many COVID-19 cases in each demographic group were ever hospitalized for their illness and how many have died from their illness. Hover over a bar to see the exact number of cases in each category and the percentage of total cases that fall in that category.

Hospitalizations / Deaths

Not Hospitalized · Hospitalization Status Unknown · Hospitalized

**SEX**

| | Hospitalized | Hospitalization Status Unknown | Not Hospitalized | Total Cases | Cases per 100,000 |
|---|---|---|---|---|---|
| Female | 1,501 | 2,325 | 19,070 | 22,896 | 1,064.6 |
| Male | 1,697 | 2,691 | 16,932 | 21,320 | 1,022.2 |
| Non-binary | 0 | 0 | 4 | 4 | |
| Refused/Unknown | 6 | 72 | 90 | 168 | |

**AGE GROUP**

| | Hospitalized | Hospitalization Status Unknown | Not Hospitalized | Total Cases | Cases per 100,000 |
|---|---|---|---|---|---|
| 9 and younger | 37 | 218 | 1,890 | 2,145 | 446.9 |
| 10 to 19 | 44 | 597 | 4,199 | 4,840 | 974.3 |
| 20 to 29 | 225 | 1,300 | 8,119 | 9,644 | 1,736.8 |
| 30 to 39 | 259 | 894 | 6,760 | 7,913 | 1,367.0 |
| 40 to 49 | 363 | 763 | 5,932 | 7,058 | 1,298.9 |
| 50 to 59 | 516 | 629 | 4,463 | 5,608 | 1,048.1 |
| 60 to 69 | 603 | 310 | 2,572 | 3,485 | 646.1 |
| 70 to 79 | 616 | 175 | 1,292 | 2,083 | 612.5 |
| 80+ | 539 | 182 | 847 | 1,568 | 935.3 |
| Refused/Unknown | 2 | 20 | 22 | 44 | |

**COVID-19 Quick Facts: Hospitalizations**

Below, you can see the percent of COVID-19 cas[illegible] who have ever been hospitalized for their illnes[illegible] broken out by sex, age group, race, and ethnicit[illegible]

| | |
|---|---|
| Female: | [illegible] |
| Male: | [illegible] |
| Ages 9 and younger: | [illegible] |
| Ages 10 to 19: | [illegible] |
| Ages 20 to 29: | [illegible] |
| Ages 30 to 39: | [illegible] |
| Ages 40 to 49: | [illegible] |
| Ages 50 to 59: | [illegible] |
| Ages 60 to 69: | [illegible] |
| Ages 70 to 79: | [illegible] |
| Ages 80+: | [illegible] |
| AI/AN*: | [illegible] |

Source: Oregon Health Authority (OHA) and the OHA Public Health Division [https://public.tableau.com/app/profile/oregon.health.authority.covid.19/viz/OregonCOVID-19CaseDemographicsandDiseaseSeverityStatewide/DemographicData](https://public.tableau.com/app/profile/oregon.health.authority.covid.19/viz/OregonCOVID-19CaseDemographicsandDiseaseSeverityStatewide/DemographicData)

#### Table I. Pennsylvania COVID-19 Cases by Age (as of October 30, 2020)

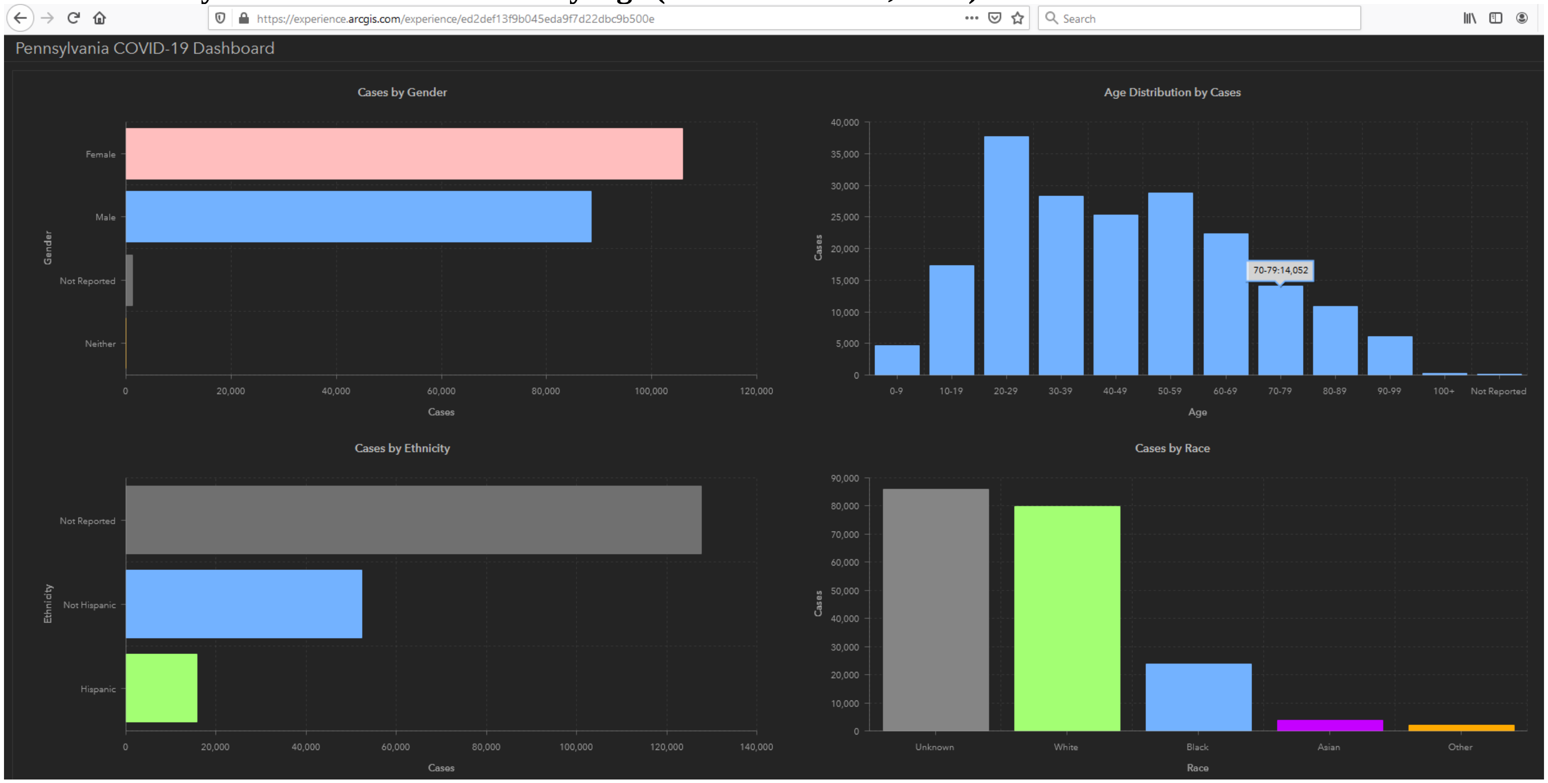


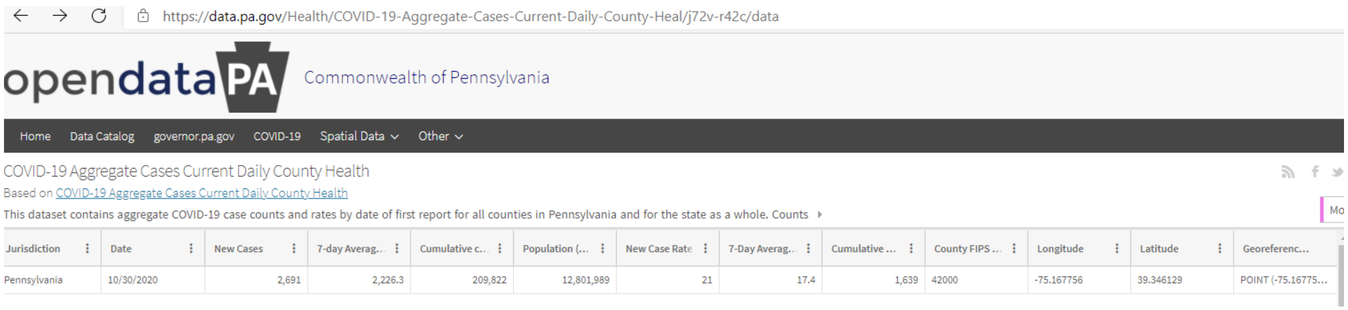

https://data.pa.gov/Health/COVID-19-Aggregate-Cases-Current-Daily-County-Heal/j72v-r42c/data

opendataPA Commonwealth of Pennsylvania

Home Data Catalog governor.pa.gov COVID-19 Spatial Data Other

COVID-19 Aggregate Cases Current Daily County Health

Based on COVID-19 Aggregate Cases Current Daily County Health

This dataset contains aggregate COVID-19 case counts and rates by date of first report for all counties in Pennsylvania and for the state as a whole. Counts

| Jurisdiction | Date | New Cases | 7-day Averag.. | Cumulative c... | Population (... | New Case Rate | 7-Day Averag... | Cumulative ... | County FIPS ... | Longitude | Latitude | Georeferenc... |
|---|---|---|---|---|---|---|---|---|---|---|---|---|
| Pennsylvania | 10/30/2020 | 2,691 | 2,226.3 | 209,822 | 12,801,989 | 21 | 17.4 | 1,639 | 42000 | -75.167756 | 39.346129 | POINT (-75.16775... |

Total cases 209,822.

Source: Pennsylvania Department of Health, https://experience.arcgis.com/experience/ed2def13f9b045eda9f7d22dbc9b500e

**Table J. South Carolina COVID-19 Cases by Age (as of Dec. 5, 2020)**

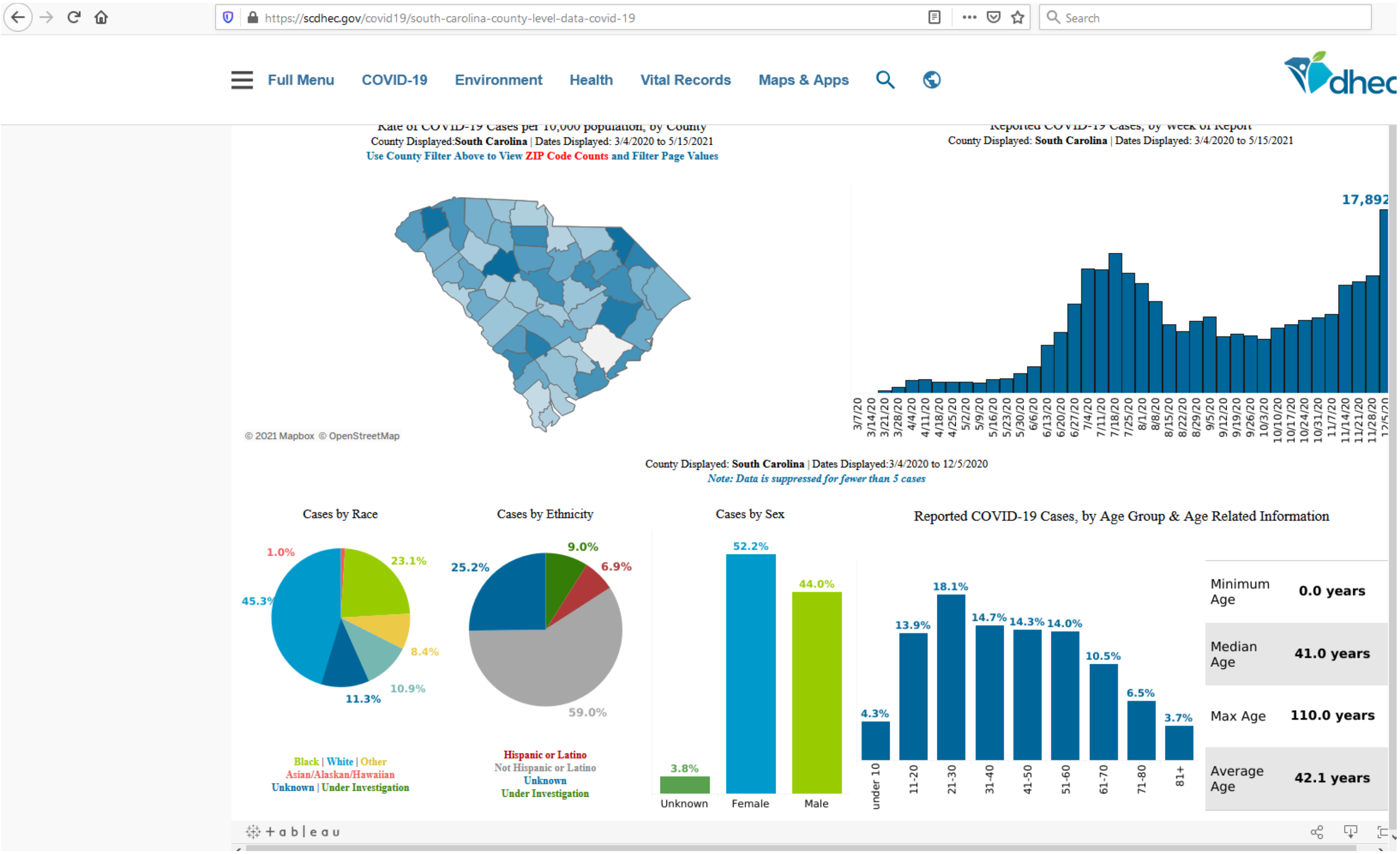


Total cases: 17,892

Source: South Carolina Department of Health and Environmental Control[https://scdhec.gov/covid19/south-carolina-county-level-data-covid-19](https://scdhec.gov/covid19/south-carolina-county-level-data-covid-19)

**Table K.  South Dakota COVID-19 Cases by Age (as of Oct. 23, 2020)**

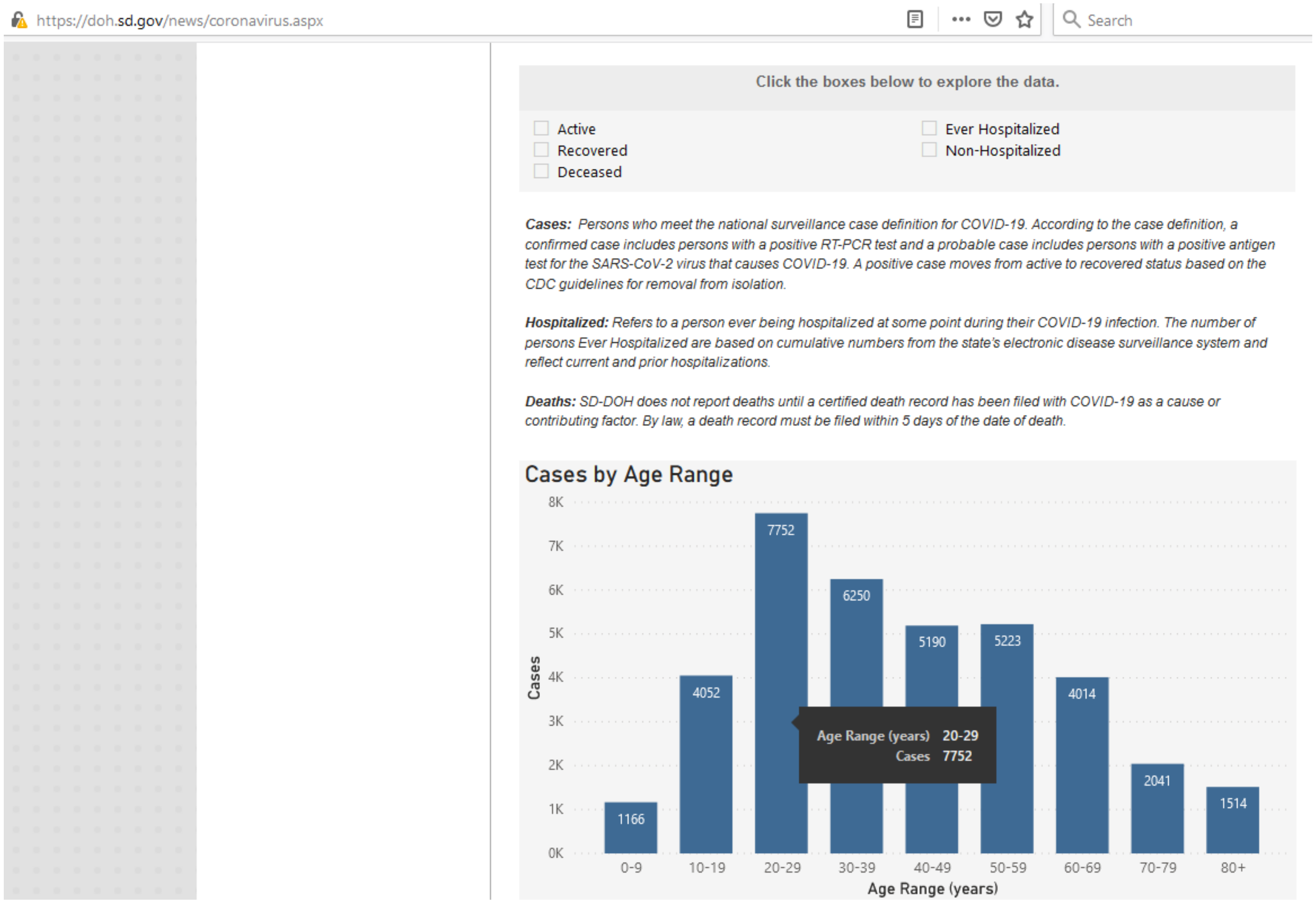


Total cases: 37,262

Source:  South Dakota Department of health
https://doh.sd.gov/news/coronavirus.aspx

**Table L. Tennessee COVID-19 Cases by Age (as of November 11, 2020)**

| Age | Total Cases |
|---|---|
| 0-10 | 14,363 |
| 11-20 | 38,925 |
| 21-30 | 57,829 |
| 31-40 | 46,794 |
| 41-50 | 43,426 |
| 51-60 | 39,243 |
| 61-70 | 27,287 |
| 71-80 | 16,316 |
| 81+ | 8,801 |
| Pending | 397 |
| Total | 293,381 |

Source: Tennessee Department of Health, "Age Dataset" in https://www.tn.gov/health/cedep/ncov/data/downloadable-datasets.html

**Table M. Wisconsin COVID-19 Cases by Age (as of Oct. 23, 2020)**

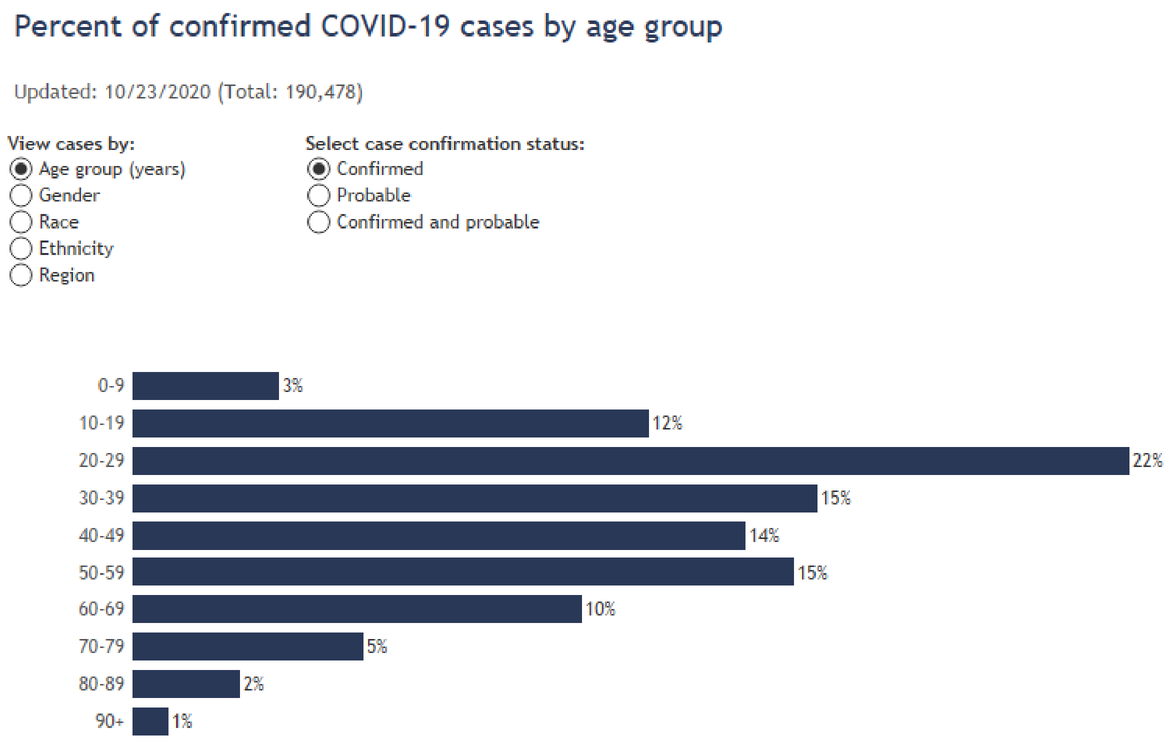


Source: Wisconsin Department of Health Services
https://www.dhs.wisconsin.gov/covid-19/cases.htm#by%20age

**Table N. Alabama COVID-19 Cases by Age (as of November 13, 2020)**

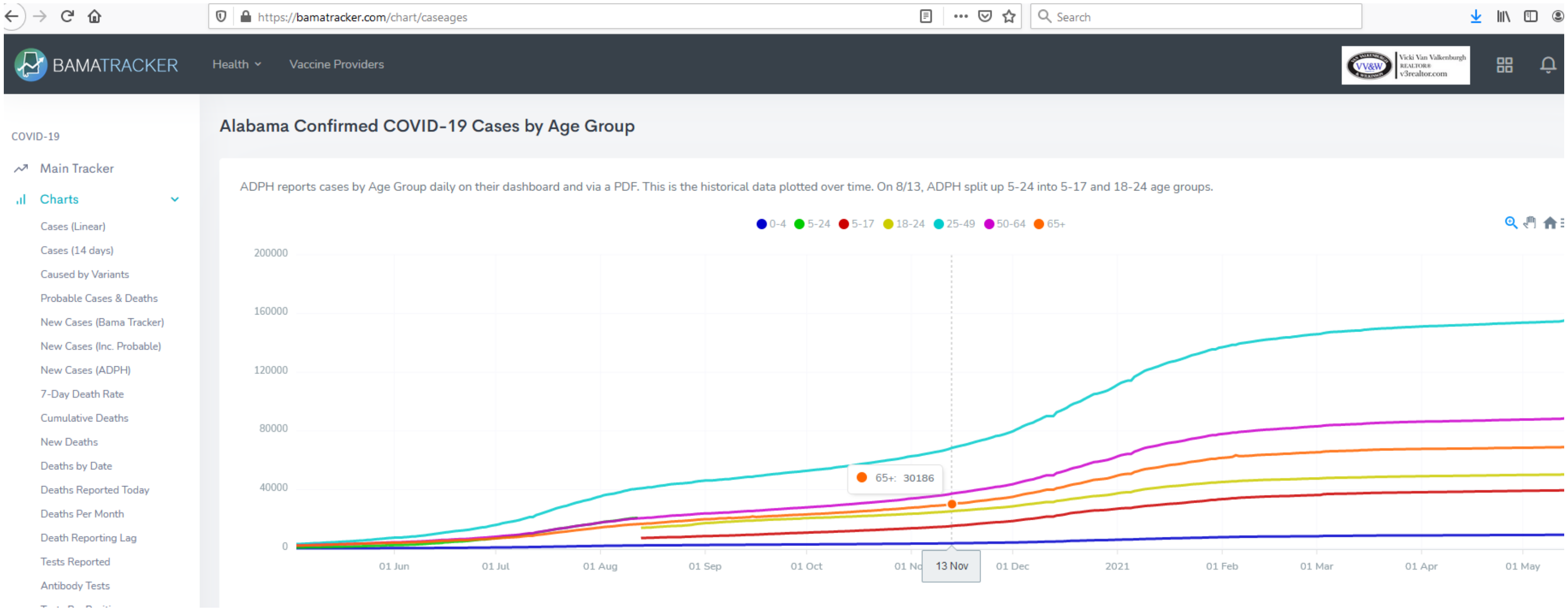


| Age | Cumulative Cases |
|---|---|
| 0-4 | 3,519 |
| 5-17 | 15,316 |
| 18-24 | 25,413 |
| 25-49 | 68,497 |
| 50-64 | 37,344 |
| 65+ | 30,186 |

Source: Alabama Department of Public Health COVID-19 Dashboard Hub
https://bamatracker.com/chart/caseages

Table O. Florida COVID-19 Cases by Age (as of Nov. 12, 2020)

### COVID-19: characteristics of 858,585 Florida resident cases

**Data through Nov 12, 2020 verified as of Nov 13, 2020 at 09:25 AM**

Data in this report are provisional and subject to change.

| Age group | Cases | | Hospitalizations | | Deaths | |
|---|---|---|---|---|---|---|
| 0-4 years | 14,429 | 2% | 336 | 1% | 0 | 0% |
| 5-14 years | 39,056 | 5% | 310 | 1% | 5 | 0% |
| 15-24 years | 140,515 | 16% | 1,376 | 3% | 32 | 0% |
| 25-34 years | 156,421 | 18% | 2,870 | 6% | 101 | 1% |
| 35-44 years | 135,609 | 16% | 4,282 | 8% | 302 | 2% |
| 45-54 years | 133,416 | 16% | 6,404 | 12% | 714 | 4% |
| 55-64 years | 111,472 | 13% | 9,001 | 17% | 1,920 | 11% |
| 65-74 years | 67,147 | 8% | 10,310 | 20% | 3,726 | 21% |
| 75-84 years | 38,283 | 4% | 9,751 | 19% | 5,096 | 29% |
| 85+ years | 21,217 | 2% | 6,895 | 13% | 5,549 | 32% |
| Unknown | 1,020 | 0% | 7 | 0% | 0 | 0% |
| **Total** | **858,585** | | **51,542** | | **17,445** | |

| Gender | Cases | |
|---|---|---|
| Female | 441,752 | 51% |
| Male | 412,389 | 48% |
| Unknown | 4,444 | 1% |
| **Total** | **858,585** | |

| Ethnicity | Cases | | Hospitalizations | | Deaths | |
|---|---|---|---|---|---|---|
| Hispanic | 254,865 | 30% | 14,431 | 28% | 4,418 | 25% |
| Non-Hispanic | 384,010 | 45% | 34,293 | 67% | 11,754 | 67% |
| Unknown | 219,710 | 26% | 2,818 | 5% | 1,273 | 7% |
| **Total** | **858,585** | | **51,542** | | **17,445** | |

| Race and ethnicity | Cases | | Hospitalizations | | Deaths | |
|---|---|---|---|---|---|---|
| White | 418,331 | 49% | 31,824 | 62% | 12,233 | 70% |
| Hispanic | 168,168 | 20% | 10,125 | 20% | 3,513 | 20% |
| Non-Hispanic | 223,614 | 26% | 20,647 | 40% | 8,124 | 47% |
| Unknown | 26,549 | 3% | 1,052 | 2% | 596 | 3% |
| Black | 125,921 | 15% | 11,771 | 23% | 3,245 | 19% |
| Hispanic | 6,239 | 1% | 493 | 1% | 128 | 1% |
| Non-Hispanic | 109,768 | 13% | 10,963 | 21% | 2,990 | 17% |
| Unknown | 9,914 | 1% | 315 | 1% | 127 | 1% |
| Other race | 117,325 | 14% | 6,293 | 12% | 1,384 | 8% |
| Hispanic | 59,750 | 7% | 3,460 | 7% | 658 | 4% |
| Non-Hispanic | 40,638 | 5% | 2,526 | 5% | 592 | 3% |
| Unknown | 16,937 | 2% | 307 | 1% | 134 | 1% |
| Unknown race | 197,008 | 23% | 1,654 | 3% | 583 | 3% |
| Hispanic | 20,708 | 2% | 353 | 1% | 119 | 1% |
| Non-Hispanic | 9,990 | 1% | 157 | 0% | 48 | 0% |
| Unknown | 166,310 | 19% | 1,144 | 2% | 416 | 2% |
| **Total** | **858,585** | | **51,542** | | **17,445** | |

**Hospitalization** counts include anyone who was hospitalized at some point during their illness. It does not reflect the number of people currently hospitalized. **Other race** includes any person with a race of American Indian/Alaskan native, Asian, native Hawaiian/Pacific Islander, or other.

Source: Florida Department of Health, htpps://Floridahealthcovid19.gov/

**Table P. Oklahoma COVID-19 Cases by Age (as of Oct. 22, 2020)**

https://coronavirus.health.ok.gov/sites/g/files/gmc786/f/2020.10.23_weekly_epi_report.pdf

#### DEMOGRAPHIC INFORMATION as of October 22, 2020

| | Cases count (%)[1] | | Deaths count (%)[1] | | Cumulative Incidence Rate[2] | Cumulative Mortality Rate[2] |
|---|---|---|---|---|---|---|
| **Oklahoma** | 112,483 | | 1221 | | 2,852.7 | 31.0 |
| **Gender** | | | | | | |
| Male | 53,835 | (47.9) | 680 | (55.7) | 2,756.5 | 34.8 |
| Female | 58,648 | (52.1) | 541 | (44.3) | 2,947.1 | 27.2 |
| **Age group** | | | | | | |
| Under 1- 4 | 2,126 | (1.9) | 0 | (0.0) | 816.3 | 0.0 |
| 5-14 | 6,392 | (5.7) | 1 | (0.1) | 1,188.3 | 0.2 |
| 15-24 | 22,499 | (20.0) | 2 | (0.2) | 4,183.3 | 0.4 |
| 25-34 | 19,221 | (17.1) | 11 | (0.9) | 3,538.4 | 2.0 |
| 35-44 | 17,855 | (15.9) | 19 | (1.6) | 3,641.0 | 3.9 |
| 45-54 | 15,456 | (13.7) | 56 | (4.6) | 3,357.7 | 12.2 |
| 55-64 | 13,080 | (11.6) | 150 | (12.3) | 2,650.8 | 30.4 |
| 65-74 | 8,698 | (7.7) | 324 | (26.5) | 2,425.2 | 90.3 |
| 75-84 | 4,771 | (4.2) | 333 | (27.3) | 2,541.7 | 177.4 |
| 85+ | 2,384 | (2.1) | 325 | (26.6) | 3,257.0 | 444.0 |
| Unknown | 1 | (0.0) | | | | |
| **Race** | | | | | | |
| American Indian or Alaska Native | 11,056 | (9.8) | 113 | (9.3) | 3,599.2 | 36.8 |
| Asian or Pacific Islander | 3,229 | (2.9) | 25 | (2.0) | 3,646.4 | 28.2 |
| Black or African American | 7,734 | (6.9) | 76 | (6.2) | 2,688.4 | 26.4 |
| Multiracial/Other | 4,435 | (3.9) | 38 | (3.1) | 1,071.9 | 9.2 |
| White | 65,252 | (58.0) | 879 | (72.0) | 2,292.8 | 30.9 |
| Unknown | 20,777 | (18.5) | 90 | (7.4) | | |
| **Ethnicity** | | | | | | |
| Hispanic or Latino | 17,450 | (15.5) | 78 | (6.4) | 4,066.9 | 18.2 |
| Not Hispanic or Latino | 71,241 | (63.3) | 994 | (81.4) | 2,027.3 | 28.3 |
| Unknown | 23,792 | (21.2) | 149 | (12.2) | | |

1. Percentages may not add up to 100 due to rounding.
2. Rate per 100,000 population

COVID-19 Weekly Epidemiology Report / October 16 – October 22, 2020 / Updated October 22, 2020 Page 11 of 23

Source: Oklahoma State Department of Health

https://coronavirus.health.ok.gov/sites/g/files/gmc786/f/2020.10.23_weekly_epi_report.pdf

**Table Q. Rhode Island COVID-19 Cases by Age (as of October 24, 2020)**

| Age | Cumulative Cases |
|---|---|
| 0-4 | 480 |
| 5-9 | 531 |
| 10-14 | 656 |
| 15-18 | 1174 |
| 19-24 | 3522 |
| 25-29 | 2655 |
| 30-39 | 4658 |
| 40-49 | 4125 |
| 50-59 | 4175 |
| 60-69 | 2880 |
| 70-79 | 1773 |
| 80+ | 2364 |
| Total cases | 28,993 |

Source: Rhode Island Department of Health (RIDOH) SalesForce COVID-19 Case Dataset
https://ri-department-of-health-covid-19-case-data-rihealth.hub.arcgis.com/#age

**Table R. Minnesota COVID-19 Cases by Age (as of Oct. 23, 2020)**

| Age Group | Number of Cases |
|---|---|
| **0-4 years** | 2,536 |
| **5-9 years** | 2,624 |
| **10-14 years** | 3,988 |
| **15-19 years** | 11,769 |
| **20-24 years** | 16,429 |
| **25-29 years** | 12,611 |
| **30-34 years** | 11,466 |
| **35-39 years** | 10,447 |
| **40-44 years** | 9,371 |
| **45-49 years** | 8,916 |
| **50-54 years** | 8,893 |
| **55-59 years** | 8,242 |
| **60-64 years** | 6,536 |
| **65-69 years** | 4,415 |
| **70-74 years** | 3,393 |
| **75-79 years** | 2,561 |
| **80-84 years** | 2,066 |
| **85-89 years** | 1,754 |
| **90-94 years** | 1,209 |
| **95-99 years** | 524 |
| **100+ years** | 95 |
| **Unknown/missing** | 18 |
| Total | 129,863 |

Source: https://www.health.state.mn.us/diseases/coronavirus/situation.html#ageg1

**Table S.  Missouri COVID-19 Cases by Age (as of Oct. 23, 2020)**

| Age | Number of Cases |
|---|---|
| 0-9 | 3054 |
| 10-19 | 13637 |
| 20 | 3596 |
| 21 | 3512 |
| 22 | 2831 |
| 23 | 2655 |
| 24 | 2403 |
| 25 | 2316 |
| 26 | 2260 |
| 27 | 2243 |
| 28 | 2181 |
| 29 | 2114 |
| 30 | 2006 |
| 31 | 1942 |
| 32 | 1893 |
| 33 | 1762 |
| 34 | 1769 |
| 35 | 1761 |
| 36 | 1659 |
| 37 | 1761 |
| 38 | 1736 |
| 39 | 1761 |
| 40 | 1752 |
| 41 | 1554 |
| 42 | 1589 |
| 43 | 1659 |
| 44 | 1579 |
| 45 | 1654 |
| 46 | 1556 |
| 47 | 1620 |
| 48 | 1678 |
| 49 | 1783 |
| 50 | 1686 |
| 51 | 1704 |
| 52 | 1616 |

| Age | Number of Cases |
|---|---|
| 53 | 1619 |
| 54 | 1579 |
| 55 | 1655 |
| 56 | 1633 |
| 57 | 1677 |
| 58 | 1619 |
| 59 | 1608 |
| 60 | 1519 |
| 61 | 1477 |
| 62 | 1361 |
| 63 | 1394 |
| 64 | 1308 |
| 65 | 1188 |
| 66 | 1100 |
| 67 | 1028 |
| 68 | 1031 |
| 69 | 976 |
| 70 | 950 |
| 71 | 886 |
| 72 | 908 |
| 73 | 844 |
| 74 | 731 |
| 75 | 707 |
| 76 | 679 |
| 77 | 746 |
| 78 | 627 |
| 79 | 574 |
| 80-84 | 2410 |
| 85-89 | 1918 |
| 90+ | 1975 |
| Unknown | 289 |

Total cases: 120,298

Source: Missouri Department of Health and Senior Services

https://health.mo.gov/living/healthcondiseases/communicable/novel-coronavirus/cases-by-age.php
